# SARS-CoV-2 UK, South African and Brazilian Variants in Karachi- Pakistan

**DOI:** 10.1101/2021.04.09.21255179

**Authors:** Adnan Khan, Muhammad Hanif, Sarosh Syed, Akhtar Ahmed, Saqib Ghazali, Rafiq Khanani

**Affiliations:** Karachi Institute of Radiotherapy and Nuclear Medicine (KIRAN), Karachi; Advanced Laboratories, Karachi; Department of Molecular Pathology, Hashmanis Group of Hospitals, Karachi; Citilab Diagnostic Center, Karachi; Global Research and Reference Labs, Karachi

## Abstract

COVID-19 pandemic has been evolving in Pakistan since the UK, South African and Brazilian variants have started surfacing which are known for increase transmissibility and can also be responsible for escape from immune responses. The gold standard to detect these variants of concern is sequencing, however routine genomic surveillance in resource limited countries like Pakistan is not always readily available. With the emergence of variants of concern and a dearth of facilities for genomic scrutiny leaves policy makers and health authorities an inconsistent and twisted image to make decisions. The inadvertent detection of B.1.1.7 by target failure because of a key deletion in spike Δ69-70 in the UK by commercially available COVID-19 PCR assay helps to understand target failures as an alternative approach to detect variants. It was ascertained further that a deletion in the ORF1a gene (ORF1a Δ3675-3677) found common in B.1.1.7, B.135 and P.1 variants of concern. The Real Time Quantitative PCR (RT-qPCR) assay for detection of emergence and spread of SARS-CoV-2 variants, by these target failures is used here. The positive samples archived in respective labs were divided in two groups used in the present study. Group I constitutes 261 positive samples out of 16964 (1.53%) collected from August till September 2020. Group II include 3501 positive samples out of 46041 (7.60%) from November 2020 till January 2021. In positive samples of group I, no variant of concern was found. A staggering difference in results was noted in group II where positivity ratio increased exponentially and the variants of concern started appearing in significant numbers (53.64% overall). This is indicative that the third wave in Pakistan is due to the importation of SARS-CoV-2 variants. This calls for measures to increase surveillance by RT-qPCR which would help authorities in decision making.

## Introduction

The global outbreak of the severe acute respiratory syndrome coronavirus-2 (SARS-CoV-2), causative agent of COVID-19 disease, has ravaged the world by surprise. As of March 28, 2021 the SARS-CoV-2 related infections have surpassed over 128,247,719 and 2,804,719 deaths globally. In Pakistan, the virus has affected more than 659,116 individuals, with more than 14,256 reported deaths till date [1]. SARS-CoV-2 has been responsible for overwhelmed health care systems, coerced shutting down of schools and gatherings, and pulling the planet into an economic recession. Despite the fact that 2020 was a challenging year, 2021 seems to be not easy with the surfacing of multiple variants of SARS-CoV-2.

One of the notorious variants of SARS-CoV-2 characterized as 501Y.V1 (lineage B.1.1.7) becomes known in the United Kingdom in December 2020, [2]. Spreading like a wildfire B.1.1.7 variant has reported to be rapidly expanding to 101 countries as of March 18, 2021 [3]. A similar pattern of staggering rise in SARS-CoV-2 related infections was observed in South Africa mainly due to the emergence of 501Y.V2 (lineage B.1351) [4]. The increased transmission rates of both the variants are attributed to the mutation in receptor binding site (N501Y) of spike protein [5]. The 501Y.V2 variant presents a possible immune escape to antibodies by having two additional mutations in the spike protein (E484K and K417N) [6]. Moreover, It was reported in a concerning development that a different set of mutations (N501Y, E484K, and K417T) started surfacing in a new 501Y.V3 (P.1) variant of SARS-CoV-2 in Brazil [7].

First cases of Covid-19 in Pakistan were reported on Feb 26, 2020 which was the beginning of the arbitrary first wave that reached its peak on June 14 with 7000 cases per day and touched the low 264 cases on August 29, 2020 with trough maintained till Oct 14, 2020 which hallmarked the beginning of so called second wave. Peak of the second wave reached on December 7 with 3795 reported new infections followed by a trough on February 16, 2021 with 747 new cases per day which was not a true trough and this decline was followed by a gradual rise in daily new cases arbitrarily labeled as the third wave (Fig 1) with its new spike of 4767 cases on March 28, 2021 [8].

**Fig 1:**
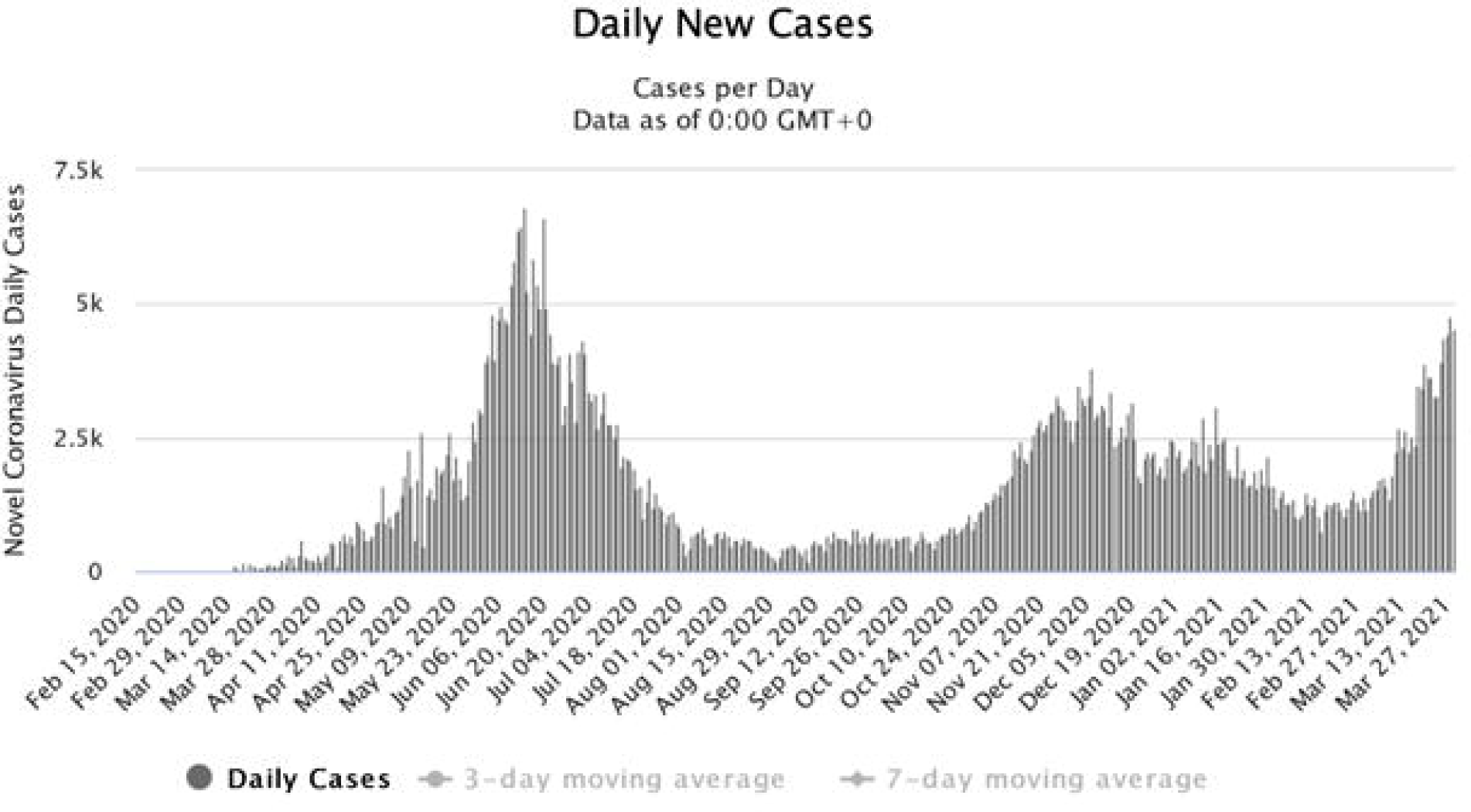
Daily record of cases of Novel Coronavirus (SARS-CoV-2). representing three peaks of SARS-CoV-2, first achieved in June 2020 followed by trough from Aug to Oct 2020. Second peak started from November and after a brief stagnant phase, the cases started to increase exponentially in March due to emergence of SARS-CoV-2 variants.

The current emergence of SARS-CoV-2 variants following the phase of genetic stability of the virus is a matter of great concern since numerous new escape variants could surface in future and can head to epidemic resurgence, as seen in other countries. Increase in transmission of the virus augment chances for the emergence of SARS-CoV-2 variants. While sequencing is the gold standard to ascertain, it cannot always be scaled right away in limited resource countries for detection of variants of concern. Therefore a multiplex qPCR assay reported earlier to detect variants of concern is used here, this can be helpful in rapid mapping of SARS-CoV-2 variants in countries like Pakistan (9).

## Materials & methods

### Ethical approval

The study was approved by Ethical Review Committee at Karachi Institute of Radiotherapy and Nuclear Medicine (KIRAN), Karachi, Pakistan.

### Sampling of SARS-CoV-2

Nasopharyngeal and oropharyngeal specimens collected for testing of SARS-CoV-2 by reverse transcription (RT) polymerase chain reaction (RT-qPCR). This was achieved in Molecular pathology section of Advanced Laboratories, Hashmanis Laboratories, and Citilab Diagnostic Centers in Karachi. The samples which came positive were archived in respective labs and were used in the present study for variant detection. Briefly, positive samples were extracted using viral Nucleic acid kit (Systaac) according to manufacturers’ instruction. The extracts were then subjected to RT-qPCR. The samples before extraction were divided in two categories, samples collected and archived from August to October 2020 (group I) and the samples collected from November 2020 to January 2021 (group II) respectively.

### Primers/ Probes

The primers/ probes set CDC_N1, Δ69/70 del, and ΔORF1a-del (Table 1) were used in the assay to detect all the three variants by targeting the Δ3675-3677 SGF deletion in the ORF1a gene (not been reported in other SARS-CoV-2 lineages) and by targeting the Δ69/70 HV deletion in the spike gene. This protocol can discriminate B.1.1.7 from B.1.351 and P.1. The 100 µM stock solutions of primers/ probes were prepared for further processing.

**Table 1:**
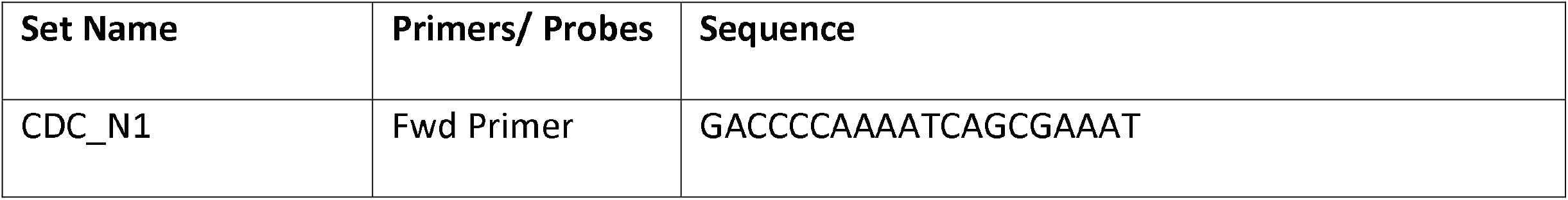
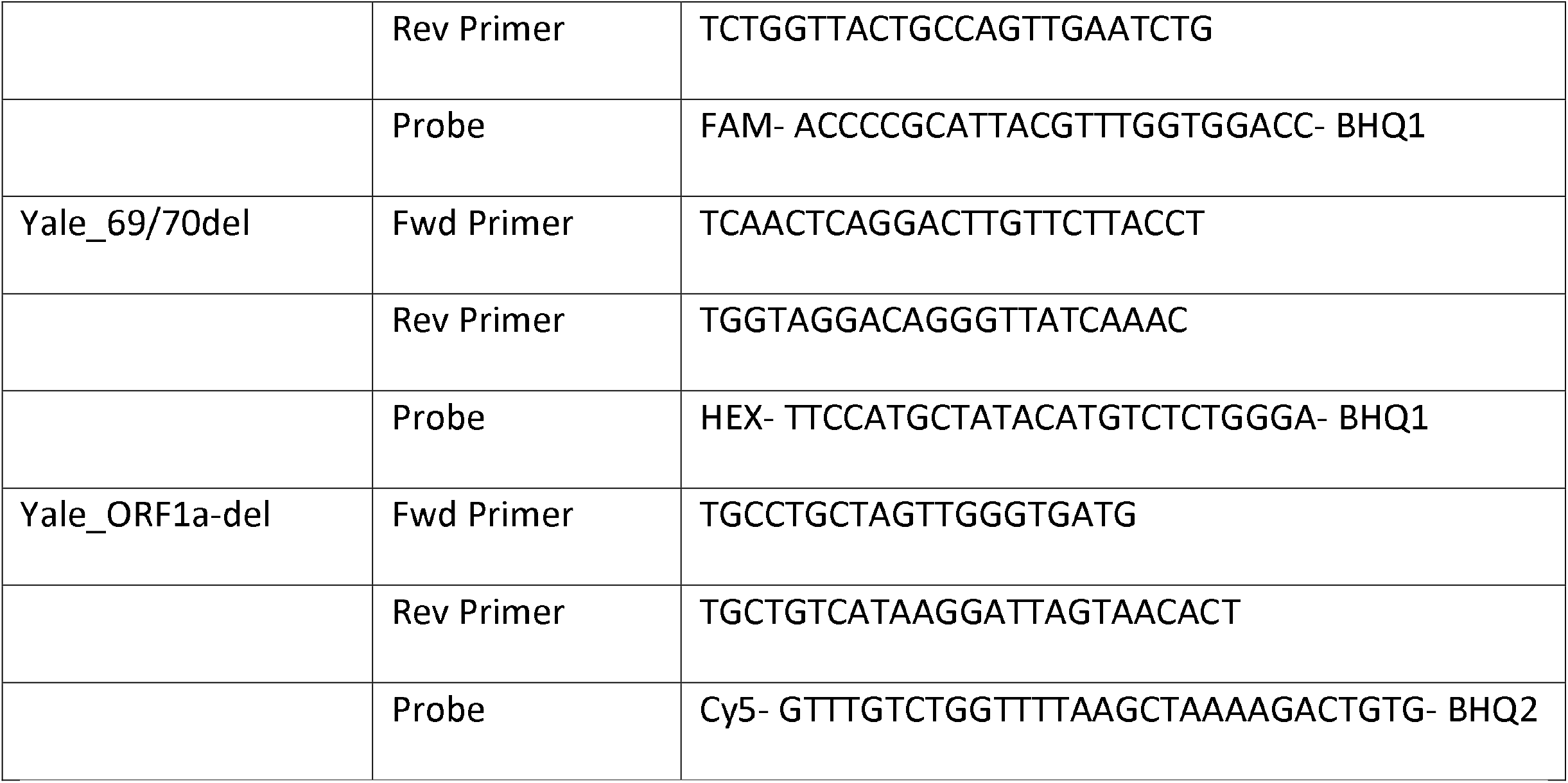
Primers/probes sequences used in multiplex RT-qPCR for detection of SARS-CoV-2 variants. Listed are three primer-probe sets used to target the neuclocapsid (N1) gene on FAM, spike Δ69-70 deletion on HEX, and ORF 1a Δ3675-3766 deletion of Cy5 fluorophores respectively.

### Multiplex RT-qPCR

The multiplex RT-qPCR assay used here was reported earlier by Vogels et al., (2021) [9]. Briefly, for PCR reaction a probe qPCR master mix along with hot start RT enzyme mix (Thermo Scientific) is used. For PCR reaction 20 µM of working stocks of primers/ probes mix was prepared using 100 µM stocks. After preparation of mix, the thermal cycler was set for FAM, HEX, and Cy5 fluorophores while the thermal cycling conditions were slightly modified for optimized results (Table 2).

**Table 2:**
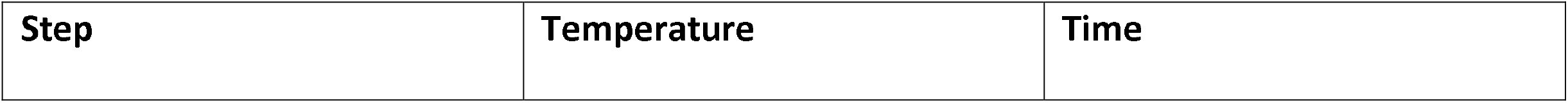
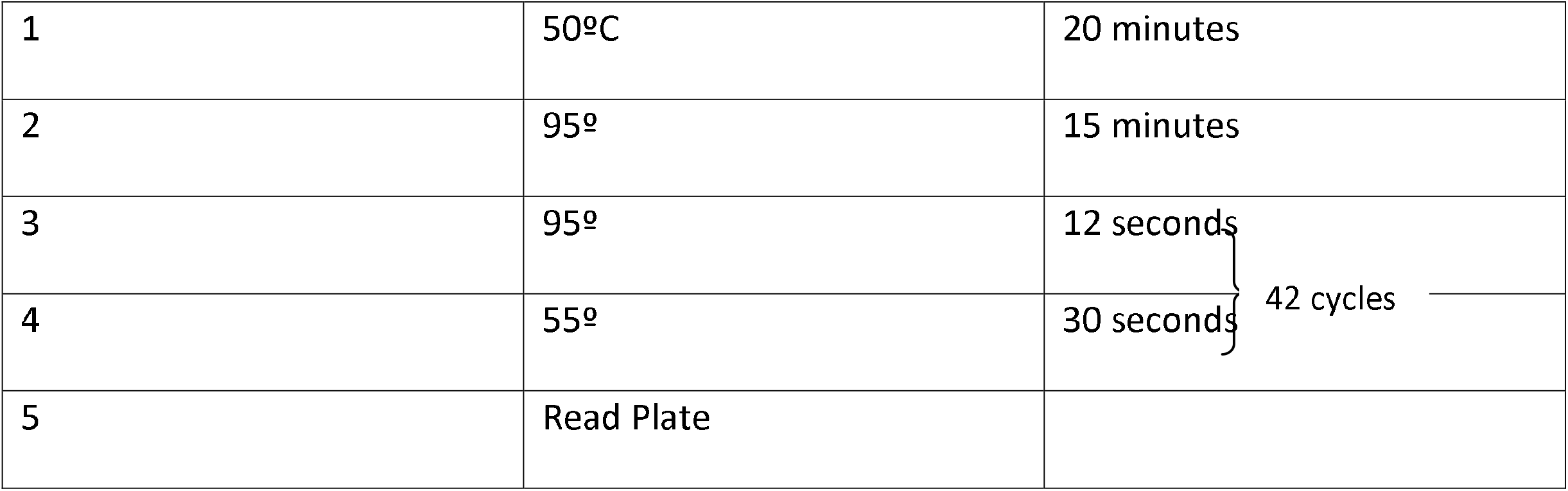
Thermal cycling programming on Bio-Rad CFX-96 for RT-qPCR detection of SARS-CoV-2 variants. Thermal cycler condition were reverse transcription (step 1), initial denaturation for 15 minutes (step 2) followed by 42 cycles of denaturation, annealing and plate read (step 3-5).

### Interpretation

All positive samples, subjected to the above mentioned RT-qPCR assay were analyzed and interpreted as exhibited in Table 3.

**Table 3:**
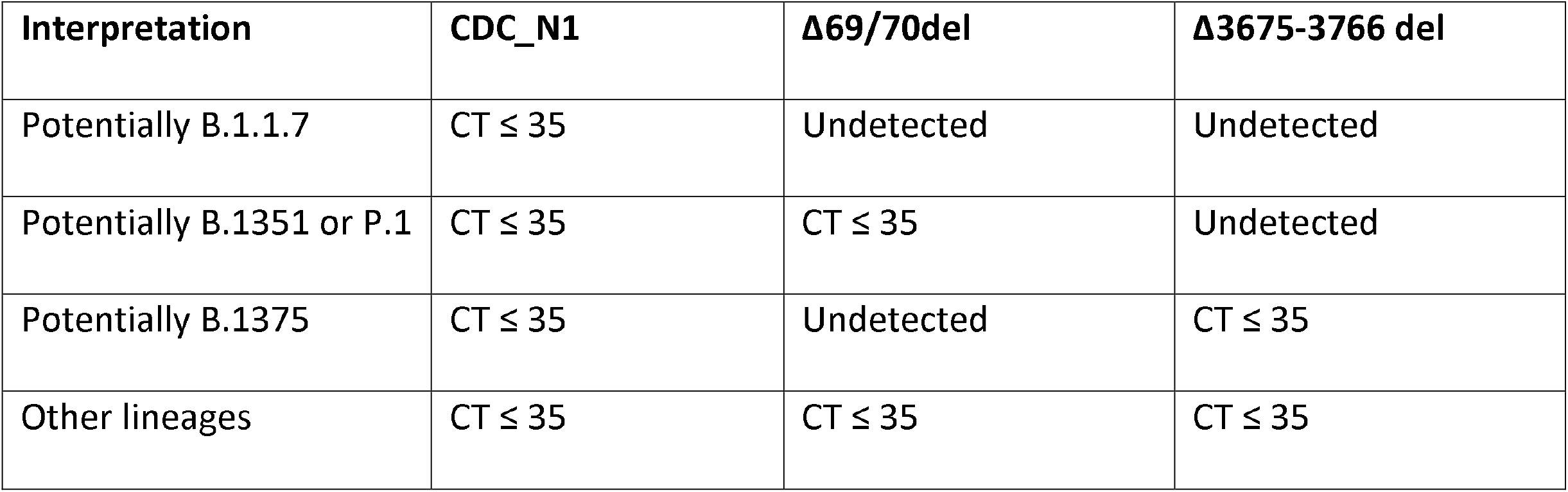
Interpretation of results of RT-qPCR assay for SARS-CoV-2 variants. Differentiation among the variants of concern is based on the target failure of spike (Δ69/70) and ORF1a (ΔΔ3675-3677) deletion.

## Results

The total tests in group I were 16964 out of which positive cases were 261(1.53%). In group II 46041 tests were performed and the positive samples were 3501(7.60%) (Table 4). Mean age of the individuals in group I was 38.01 (±16.403), and in group II 34.05 (±15.219) (Table 5). Only 58 out of 261 positive samples in group I were female (22.22%) and in group II 950 out of 3501 were female (27.01%).

**Table 4:** Scenario of SARS-CoV-2 B.1.1.7, B.1.351, and P.1 variants of concern in Karachi, Pakistan. Describes the total number of samples in group I and group II, percentage of positive samples in both groups and the occurrence of variants of concern in both groups. Data in group I indicates the low positivity ratio and non existence of variants of concern. In group II positivity ratio increased significantly and the ratio of variants of concern is very high and almost semi of the other lineages.

|  | Period selected for analysis | Total #of samples received for RT-qPCR testing | Total # of PCR positive samples | Percentage of PCR positive samples | Scenario of SARS-CoV-2 B.1.1.7, B.1.351, and P.1 variants of concern in Karachi |  |  |
| --- | --- | --- | --- | --- | --- | --- | --- |
|  |  |  |  |  | B.1.1.7 | B.1.351 & P.1 | Other lineages |
|  | August | 4638 | 90 | 1.94% | 0 | 0 | 90 |
|  | September | 5833 | 69 | 1.182% | 0 | 0 | 69 |
|  | October | 6493 | 102 | 1.57% | 0 | 0 | 102 |
| Group I |  | 16964 | 261 | 1.538% | 0 | 0 | 261 |
|  | November | 9233 | 610 | 6.60% | 164(26.88%) | 68(11.14%) | 378(61.96%) |
|  | December | 19299 | 1682 | 8.71% | 474(28.18%) | 484(28.77%) | 724(43.04%) |
|  | January | 17509 | 1209 | 6.90% | 326(26.96%) | 362(29.94%) | 521(43.09%) |
| Group II |  | 46041 | 3501 | 7.60% | 944(26.96%) | 934(26.67%) | 1623(46.3%) |

**Table 5:** Demography of sample population and the variants of concern in Karachi, Pakistan. Describe the gender, age and positivity ratio in group I and group II.

|  | Total #of samples for RT-qPCR testing | Total # of PCR positive samples | Positivity percentage | Male to Female ratio |  | Mean age |
| --- | --- | --- | --- | --- | --- | --- |
|  |  |  |  | Male | Female |  |
| Group I | 16964 | 261 | 1.538% | 77.85% | 22.15% | 38.01<br>(±16.403) |
| Group II | 46041 | 3501 | 7.60% | 72.99 | 27.01% | 34.05<br>(±15.219) |

SARS-CoV-2 variants of concerned were detected by target failures using a multiplex RT-qPCR. A descriptive RT-qPCR amplification graph is shown in Fig 2. Group I was screened and none of variants of concern were found to be present (All targets amplified). In group II significant difference noted from group I, the results showed a total of 944 (26.96%) showed double target failure of Δ69-70 as well as Δ3675-3766 del which is indicative of B.1.1.7 variant of concern (UK variant). A rather significant number 934 (26.67%) samples exhibited single target failure Δ3675-3766-del implied presence of B.135 and P.1 variant of concern. While 1623 (46.30%) samples did not show any failure of target and represent other lineages.

**Fig 2.**
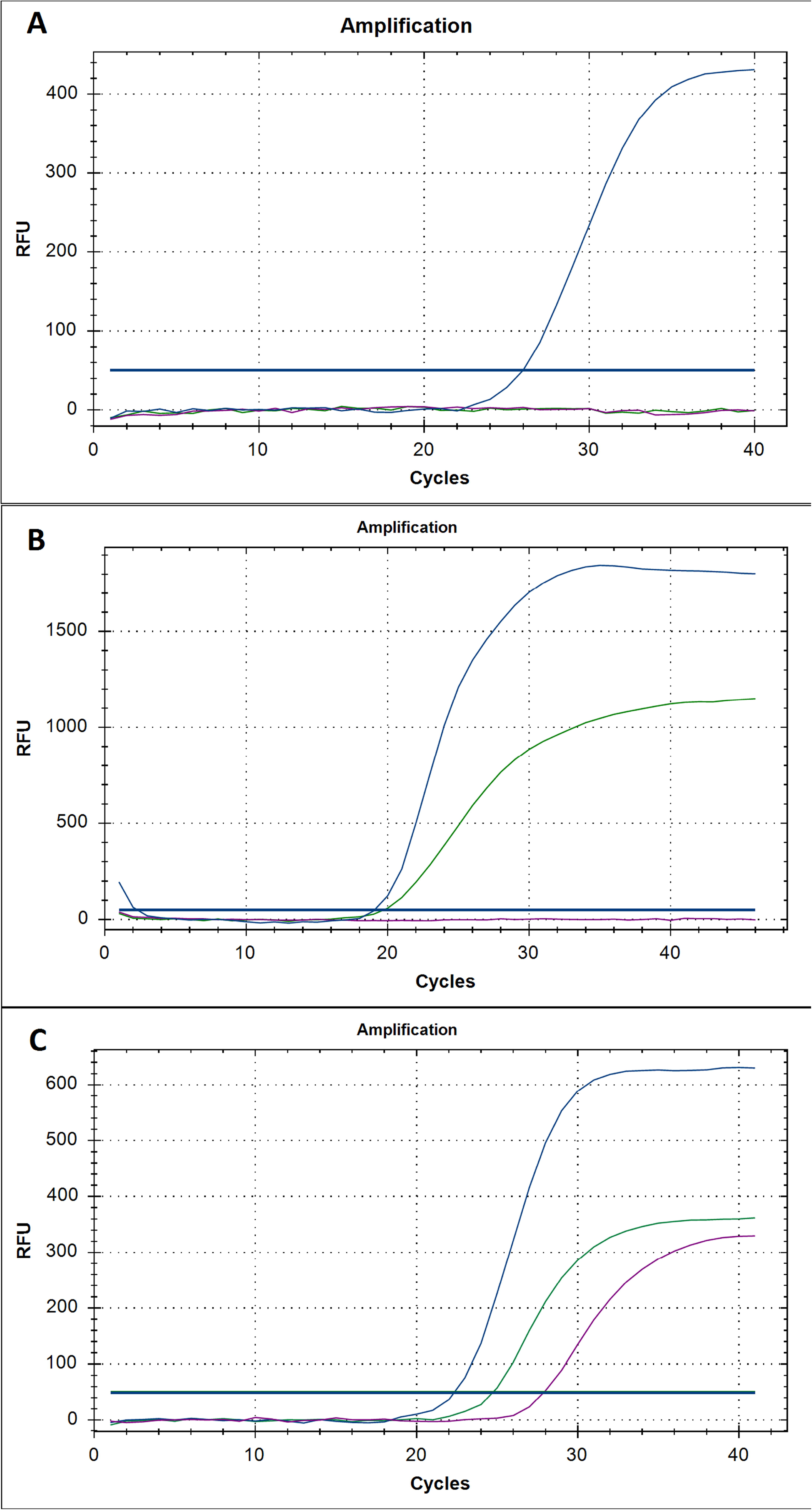
RT-qPCR graphical representation of ORF1a and spike target failure to discriminate B.1.1.7, B.135, P.1 and other lineages of SARS-CoV-2. (A) exhibit double target failure by ORF1a Δ3675-3677 and spike Δ60-70 deletions, indicative of potential B.1.1.7. (B) shows potential B.135 and P.1 variants of SARS-CoV-2 by ORF 1a target failure. (C) All three targets of the multiplex RT-qPCR demonstrate no target failure and lineages other than variants of concern.

## Discussion

The PCR tests for SARS-CoV-2 detection in UK during September 2020 have shown unusual SARS-CoV-2 strain that prevents amplification of spike gene, also known as spike gene target failure (SGTF), in a commercially available kit [10]. The failure to detect spike gene is attributed to a Δ69/70 HV deletion as in the lineage 501.YV1 (B.1.1.7) or commonly known as the UK variant. Moreover, the P.1. lineage in Brazil and the B.1.351 lineage described in South Africa share a common set of mutations ORF1a deletion 3675-3677 SGF [11]. Therefore, a sample with Δ69/70 HV deletion is not enough for discriminative detection of B.1.1.7 from other variants of concern i.e501.YV2 and 501.YV3 (B.1351; South African and P.1; Brazilian variant), which does not possess Δ69/70 HV deletion. Therefore, a multiplex PCR assay reported earlier targeting Δ3675-3677 SGF deletion in ORF 1a gene along with Δ69/70 HV deletion is used in the present study to discriminate B.1.1.7 from B.1351 and P.1 variants of SARS-CoV-2[9]. We have used CDC_N1 primer and probe as a control to ensure that target failures were due to these mutations. In resource limited countries like Pakistan the present study can therefore be helpful to rapidly scale up the prediction of SARS-CoV-2 variants.

These SARS-CoV-2 variants (B.1.1.7, B.1.351 and P.1) are the cause of concern globally not only because of their ability to transmit rapidly but also because of antigenic changes due to multiple mutations in spike protein region unfavorable to monoclonal antibody (mAB) therapies and protection by vaccines [7, 12]. The variant P.1 or 501Y.V3 is spreading at a rapid pace in Brazil and spreading globally. N501Y is the common mutation shared between these three SARS-CoV-2 lineages of concern. This particular mutation may result in augmented binding to ACE231 which exhibit no significant escape value from emergency use authorized mAB therapies and vaccines [13]. However, E484K mutation present in B.135 and P.1 variants has been attributed for impairing many mABs and vaccines under development and also responsible for re-infection [10]. This threat has been augmented by the recent observation from the Novavax vaccine trial in South Africa which showed that exposure to B.135 in placebo recipients’ exhibit to be un-protective against prior SARS-CoV-2 infection [12-14]. Even in Brazil the same mutation in P.1 is reported to be responsible for spreading of the virus in a population that was already 76% seropositive due to past infection [14]. This highlights the importance of this study of mapping viral variants by RT-qPCR which could be adopted by limited resource countries worldwide. This mapping would be helpful in effective development of therapies and vaccines targeting antigenically distinct epitopes.

We demonstrate the presence of UK (B.1.1.7; 27.04%) and South African and Brazilian (B.135 and P.1; 29.94%) variants of SARS-CoV-2 from the positive samples collected in group II. the present study positive samples archived before November 2020,i.e in group I, did not exhibit variants of concern. Noteworthy, in group I (Aug to Oct 2020) the positivity ratio was 1.538% which later increased to 7.60% in group II (Nov 2020 to Jan 2021) which clearly indicate the importation of cases from countries where these viral strains were spreading before and during November 2020. The spread of pandemic globally provides opportunity for variable strain types to thrive. One of the reasons of this spread could be the unmonitored international flights and no quarantine on arrival in Pakistan from any country, which lead to a rapid introduction of variants of concern in Pakistan [15-16]. Pakistan is in second wave which could not be truncated due to variants entry which prevented decline to base line following the first wave and second peak of second wave is labeled as 3^rd^ wave affecting Prime minister, First Lady and President of Pakistan. Not only the notables of the country have been affected in this notorious second wave, but our results also revealed that positivity ratio in female has significantly increased from 22.15% to 27.01%. This clearly indicates that variants are more transmittable.

At present, the significance of the surveillance of variants has increased globally as the vaccination jabs have started and countries are in race to vaccinate. Limited resources countries are lagging behind and with the emergence of these variants in these countries it is of high possibility that an escape variant could evolve which could hinder the vaccination process globally. The limited resource countries might act as the breeding ground for the variants of the virus that could be detrimental to global efforts for ending the pandemic.

The striking part of our study is the presence of variant with Δ3675-3677 SGF deletion in ORF 1a gene which implied the presence of South African and Brazilian variants in Pakistan. The most detrimental part of the present study is that South African and Brazilian variants are comparable in numbers in December 2020 and January 2021 as compared to its counterpart UK variant, while SARS-CoV-2 with lineages other than the variants of concern are less in numbers in positive samples collected during and after November 2020. The emergence of the UK, South African and Brazilian variants calls for strict measures to curb the spread of SARS-CoV-2. The lucidity in this situation of darkness can only be attained by arrangement of non-pharmaceutical interventions and scaling up-of vaccination program, which could lead to fewer infections, in turn leads to fewer variants. Both strategies are important until population immunity is achieved globally.

The current pandemic should nudge the developed countries that infectious diseases have a profound impact on lives and economies and rapid distribution and implementation of useful vaccines against these infections should remain priority. The cooperation of the globe to make certain the justness and responsiveness to local contexts is necessary on the complex trail forward to ending the COVID-19 pandemic.

## Data Availability

Data is fully available without restrictions

## Acknowledgements

Authors acknowledge support from IT departments of Advanced Laboratories, Hashmanis group of hospital and Citilab diagnostic center for their support in providing SARS-CoV-2 data.

